# Psychosis-linked Symptoms and Structural Brain Patterns in Cognitive Subgroups among Familial High-Risk Children in the ABCD Study

**DOI:** 10.1101/2025.01.31.25321486

**Authors:** Halide Bilge Türközer, Nicole R. Karcher, Jacqueline Clauss, Merete Nordentoft, Kathryn E. Lewandowski, Joshua L. Roffman, Dost Öngür

## Abstract

**Objective:** Children at familial high risk for psychosis (FHR) are at substantially increased risk for psychotic disorders and other serious mental illnesses. Identifying risk subgroups within FHR youth may enhance prediction models to identify children at greatest risk for potential intervention. This study investigated psychosis-linked symptoms and structural brain patterns in neurocognitive subgroups among FHR children in the Adolescent Brain Cognitive Development (ABCD) Study using baseline, 2-year, and 4-year follow-up data.

**Methods:** Among children with first- and second-degree family history of psychosis, neurocognitive subgroups were defined using NIH Toolbox Cognitive Battery baseline age-corrected total scores: children with low (FHR-LC, 0-33%, n=234), moderate (FHR-MC, 33-66%, n=261), and high (FHR-HC, 66-100%, n=277) cognitive performance. Psychiatric symptoms were assessed using Prodromal Questionnaire-Brief Child Version (PQ-BC) and Childhood Behavior Checklist (CBCL). Regional vulnerability indices (SSD-RVIs), which quantify the similarity of participants’ structural brain patterns to the patterns found in adults with schizophrenia spectrum disorders, were calculated using cortical thickness measures following rigorous quality control.

**Results:** At baseline, FHR-LC had significantly higher PQ-BC and CBCL scores, and trend-level higher SSD-RVIs compared to FHR-HC. Longitudinally, PQ-BC and CBCL scores decreased with age across all FHR participants, while SSD-RVIs remained stable. No longitudinal cognitive subgroup-by-age interactions were observed, indicating that subgroup differences persisted over time.

**Conclusion:** Children at FHR who have concurrent poor cognitive performance exhibit elevated and stable clinical and imaging psychosis risk markers. This suggests that they may represent a risk subgroup with elevated vulnerability, presenting an opportunity for early identification and intervention.

## INTRODUCTION

Identifying risk groups for psychotic disorders more accurately has been one of the major challenges our field faces, posing a bottleneck for evidence-based preventative interventions. Youth at familial high risk (FHR) for psychosis has received less attention than other subgroups in mainstream psychosis prevention paradigms and clinical services. The leading paradigm for psychosis risk has been the clinical high-risk (CHR) framework, which identifies short-term risk in youth largely based on clinical presentation and help-seeking status. However, alternative approaches to the CHR model, distinct yet complementary, could accelerate the progress towards primary prevention of psychotic disorders. FHR youth not only exhibit significantly higher rates of psychotic disorders (∼12%) compared to the general population, akin to transition rates in CHR (∼14%), but also nearly half of them develop a psychiatric disorder by the age of 20 (1). In addition to familial risk, they are at an increased likelihood of exposure to environmental risks, including childhood adversities and cannabis use (2–4). Despite their high-risk status marked by familial and environmental risks, there is a gap of knowledge about the biological heterogeneity and risk subgroups within FHR youth. This critical gap hinders the development of outcome prediction models for FHR youth and the potential for effective preventative interventions.

Although there is limited information on the biological heterogeneity within FHR, biological subgroups within FHR have been identified and may inform higher accuracy prediction models for psychosis in this risk group (5–7). In a prior study, we investigated biomarker profiles in risk subgroups among clinically unaffected first-degree relatives of individuals with psychotic disorders in the Bipolar and Schizophrenia Network for Intermediate Phenotypes (BSNIP) dataset (5). In the BSNIP dataset, unaffected FHR individuals (U-FHR) who were younger than age 30, therefore still within the typical age window for developing psychosis, had worse performance in cognitive control tasks (i.e. antisaccade error and stop-signal tasks) compared to U-FHR older than age 30 and age-matched healthy controls (5). This indicates that FHR youth who demonstrate cognitive control deficits may leave the unaffected FHR pool after age 30, possibly via conversion to psychosis. Recent findings from a prospective longitudinal cohort study, the Danish High Risk and Resilience Study, support these findings. This study showed distinct neurocognitive subgroups among FHR children, which can be detected as early as age 7 (6,7). The impaired subgroup shows higher psychopathology, poorer function, and elevated polygenic risk scores for psychosis. These findings showcase that the cognitively impaired FHR subgroup constitutes a clearly defined, easily identifiable early risk status marked by heritable, clinical, and cognitive characteristics that align with developmental models of psychotic disorders. However, whether the cognitively impaired FHR subgroup is at a particularly high risk for developing psychosis has not been assessed in longitudinal studies and this presents a critical gap in the field.

Identifying risk subgroups within FHR youth that offer improved predictive accuracy may pave the way for novel preventative interventions targeting these subgroups while the brain is still highly plastic. The current study aimed to assess psychotic-like experiences and psychosis-linked structural brain patterns in neurocognitive subgroups among FHR children in the Adolescent Brain Cognitive Development (ABCD) Study at baseline and longitudinally using the 2-year and 4-year follow-up data. To assess psychosis-linked brain patterns, we used the recently developed Regional Vulnerability Indices (RVIs). RVIs quantify the similarity of participants’ structural neuroimaging measures to the expected patterns found in adult samples with schizophrenia-spectrum disorders (SSD-RVIs) based on findings from the Enhancing Neuro Imaging Genetics Meta Analyses (ENIGMA) consortium (8). Previous work on RVIs demonstrated that children and adolescents with clinical, familial, and environmental risk factors for psychosis exhibit deficit patterns similar to those observed in adults with psychotic disorders (8,9,10). In this study, our primary hypothesis was that FHR youth with low neurocognitive functioning (FHR-LC) would demonstrate more severe psychotic-like experiences (PLE) and general psychopathology at baseline, with a steeper increase of symptoms during the follow-up period as the psychosis risk window approaches, compared to the FHR youth with high cognitive functioning (FHR-HC). We hypothesized that FHR-LC would have structural brain patterns more similar to those observed in adults with psychotic disorders, evidenced by higher SSD-RVI scores, compared to the FHR-HC group. We also predicted that structural brain patterns in FHR-LC would become increasingly more similar to those seen in psychotic disorders during the follow-up period, evidenced by a steeper increase in SSD-RVI scores, compared to the FHR-HC group.

## METHODS

The ABCD Study is a multi-center study tracking 9-10-years-olds recruited from 21 research sites across the United States. Our study included data from baseline (N= 11,868), 2-year follow-up (N= 10,973), and available data from 4-year follow-up (N=4,754), which has been partially released. We identified familial high risk for psychosis (FHR) youth using first- and second-degree family history of psychosis recorded in the ABCD dataset. Family history of psychosis was defined based on parent or primary caregiver reports of relatives seeing visions or hearing voices, or thinking of people were spying on them or plotting against them. Siblings or cousins were excluded from family history conclusion given their likely young age for a diagnosis of a psychotic disorder. 914 FHR participants were identified in the baseline ABCD dataset. Family Loading Score for psychosis was calculated by summing the expected degree of genetic sharing with the affected relative(s), using the coefficients 0.5 for an affected parent and 0.25 for an affected grandparent, aunt, or uncle(10). For example, if a participant has a parent and an uncle with psychosis, their family loading score for psychosis is 0.75 (0.5 + 0.25).

NIH Toolbox Cognition Battery® (NIHTBX), which include measures of executive function, attention, memory, language, and processing speed, was used to assess cognitive functioning. Prodromal Questionnaire-Brief Child Version (PQ-BC) was used to examine self-report of psychotic-like experiences (11,12). Childhood Behavior Checklist (CBCL) was used to assess parent report of psychopathology (13). PQ-BC total severity scores and standardized CBCL total scores (t scores) were included in the analyses.

### Structural MRI analysis

T1 weighted structural images were acquired on 3T Siemens, Philips, or GE scanners as described by Casey et al.(14). Minimally processed T1 volumes were obtained from the NIMH Data Archive and reprocessed using FreeSurfer version 7.1. Using automated segmentation (Desikan-Killiany atlas), cortical thickness of 68 regions of interest were quantified. Rigorous visual quality control was performed by a trained rater. Imaging analysis and quality control methods are described in detail by Elyounssi et al.(15). Participants with poor quality structural data (surface hole number, SHN > 62.5) were excluded (N=1213 in the combined BL, Y2, Y4 datasets).

### Schizophrenia Spectrum Disorder Regional Vulnerability Indices (SSD RVIs)

We calculated cortical SSD-RVIs using the method previously described by Kochunov et al. (10). Cortical thickness (CT) measures from the 34 DK Atlas regions were averaged across the two hemispheres to obtain mean CT values for each region. Covariates (age, sex, race and ethnicity, and intracranial volume) were then regressed out using linear regression. Unstandardized residuals were z transformed. For each participant, an SSD-RVI was calculated as a Pearson correlation coefficient between the 34 region-wise z values and their corresponding effect sizes for schizophrenia-control group differences reported by ENIGMA (16). Three outliers were identified based on the > 1.5 interquartile range (IQR) rule and excluded (two participants in the Y2 and one in the Y4 dataset). The Kolomogorov-Smirnov normality test showed that SSD-RVI values were normally distributed in the 772 FHR participants included in the final analysis (D = 0.017, p = 0.982).

### Statistical Analyses

After excluding participants with poor quality structural MRI data, outlier SSD-RVI values, and without cognitive data, 772 FHR participants with baseline data, 515 with Y2 data, and 200 with Y4 data were included in the final analysis. Prior studies that examined data-driven subgroups among FHR children showed three neurocognitive subgroups that align with severity levels (severely impaired, mildly impaired, average/above average)(7,17). For this reason, in this study, we used a percentile-based approach for grouping which is more practical for clinical translation. FHR participants were grouped into three cognitive subgroups based on the baseline NIHTB age-corrected total composite scores: The FHR group with low cognitive performance (FHR-LC, 0-33%, n=234), moderate cognitive performance (FHR-MC, 33-66%, n=261), and high cognitive performance (FHR-HC, 66-100%, n=277). Age-corrected total composite scores were not available in Y2 and Y4 data. Please see Supplementary Material for analyses examining cognitive subtest scores over time across the three cognitive subgroups (**Figure S1**).

We utilized linear mixed-effects models using the lme4 package in R to examine group differences and trajectories. In the baseline dataset, we assessed differences in PQ-BC, CBCL, and SSD-RVI scores across the three cognitive subgroups (fixed factor). For longitudinal analyses, we first evaluated the fixed effect of age in PQ-BC, CBCL, and SSD-RVI scores in the whole FHR group to assess for their change with age over time. We then examined the interaction between age and cognitive subgroup membership to examine group differences in trajectories over time. For analyses involving repeated measures across time, Subject ID was added as a random factor.

All models included family ID and site as random effects to account for clustering and site-specific effects. Covariates varied by model: PQ-BC models included age (in the baseline analysis), sex, and race-ethnicity; CBCL models included race-ethnicity only, as standardized T scores are already adjusted for age and sex. SSD-RVI models did not include additional covariates, as age, sex, race-ethnicity, and intracranial volume had been regressed out in the creation of RVI scores.

The false discovery rate (FDR) approach was used to correct for multiple testing (*q* value). FDR corrections were applied separately for the baseline and longitudinal analyses. Statistical significance was set at *q* = 0.05. For the baseline analyses, FDR correction was performed for 9 comparisons (3 subgroup comparisons x 3 outcome measures). For the longitudinal effect of age in the whole FHR group, FDR correction was performed for 3 comparisons (fixed effect of age x 3 outcome measures). For the interaction between age and cognitive subgroups longitudinally, FDR correction was performed for 9 comparisons (age*group interaction for three subgroups x 3 outcome measures).

Following the analyses on the whole dataset, we conducted a sensitivity analysis by performing a complete case analysis, including only participants with complete BL, Y2, an Y4 data (N=175, n_FHR-LC_=42, n_FHR-MC_=48, n_FHR-HC_=85) using the same methods described above. To assess the robustness of findings, we compared results from the complete case analysis with those obtained using the entire dataset.

## RESULTS

Baseline demographics and clinical/cognitive measures are presented in **Table 1**. FHR-LC and FHR-MC groups were younger than the FHR-HC group (one-way ANOVA, p<0.001). Race and ethnicity distributions were also different across the three groups (chi-square test, p<0.001), with all three groups significantly differing from each other. Family Loading Scores did not differ across the three groups (one-way ANOVA, *p* = 0.1) (**Table 1**).

**Table 1.** Demographic and clinical characteristics of cognitive subgroups among FHR children.

|  | FHR-LC<br>(N=234) | FHR-MC<br>(N=261) | FHR-HC<br>(N=277) | Statistics |
| --- | --- | --- | --- | --- |
| Age (Mean, years) | 9.80 | 9.89 | 10 | p=0.002* |
| Age (SD) | 0.59 | 0.62 | 0.63 |  |
| Sex |  |  |  | p=0.22 |
| Female | 130 | 131 | 133 |  |
| Male | 104 | 130 | 144 |  |
| Gender |  |  |  | p=0.22 |
| Female | 130 | 131 | 133 |  |
| Male | 104 | 130 | 144 |  |
| Race and Ethnicity |  |  |  | p<0.001 |
| White | 65 | 114 | 167 |  |
| Black | 75 | 50 | 17 |  |
| Hispanic | 57 | 61 | 51 |  |
| Asian | 1 | 2 | 8 |  |
| Other | 36 | 34 | 34 |  |
| Family Loading Score (Mean) | 0.38 | 0.36 | 0.35 | p=0.1 |
| Family Loading Score (SD) | 0.20 | 0.19 | 0.16 |  |
| CBCL Total t score (Mean) | 51.21 | 49.23 | 47.48 |  |
| CBCL Total t score (SD) | 12.15 | 12.59 | 11.47 |  |
| PQB Total Severity Score (Mean) | 10.80 | 8.43 | 5.47 |  |
| PQB Total Severity Score (SD) | 15.21 | 13.93 | 9.34 |  |
| NIHTB Total Composite Score (Mean) | 79.97 | 96.99 | 117.86 |  |
| NIHTB Total Composite Score (SD) | 6.98 | 4.34 | 10.43 |  |

FHR-LC: Family High Risk – Low Cognition, FHR-MC: Family High Risk – Moderate Cognition, FHR-HC: Family High Risk – High Cognition; CBCL: Childhood Behavior Checklist; PQ-BC: Prodromal Questionnaire-Brief Child Version (PQ-BC); \*age: a post-hoc analysis shows FHR-HC > FHR-LC (p<0.001), FHR-HC > FHR-MC (p=0.03).

### Comparisons of baseline measures across the cognitive subgroups

At baseline, FHR-LC had significantly higher PQ-BC severity scores compared to the FHR-HC group (β = 4.457, 95% CI [2.052,6.862], *t* = 3.63, *q* = 0.002, *d* = 0.41) (Figure 1A). FHR-LC also had higher CBCL total t-scores compared to both the FHR-MC (β = 2.606, 95% CI [0.578, 4.634], *t* = 2.52, *q* = 0.036, *d* = 0.34) and FHR-HC groups (β = 3.957, 95% CI [1.802,6.112], *t* = 3.60, *q* = 0.002, *d* = 0.52) (Figure 1B). FHR-LC had higher SSD-RVI’s compared to FHR-HC, however this difference was at trend-level significance following FDR correction (β = 0.039, 95% CI [0.0003, 0.078], *t* = 1.97, *p* = 0.048, *q* = 0.088, *d* = 0.21) (Figure 1C).

**Figure 1.**
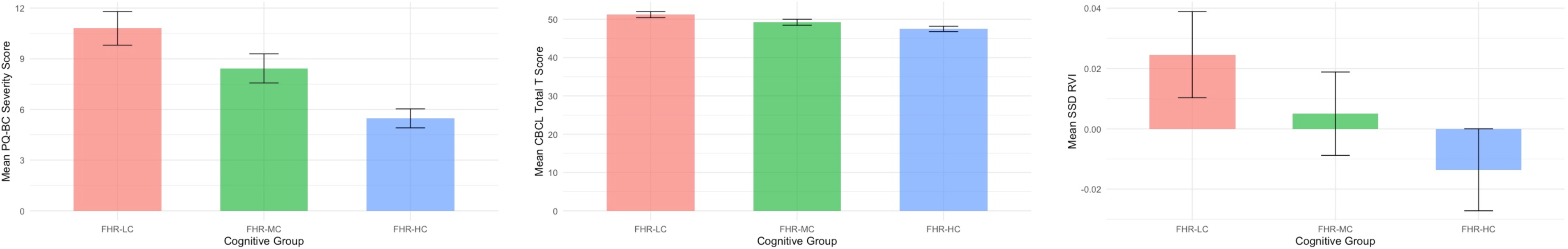
Bar graphs of mean Prodromal Questionnaire-Brief Child Version (PQ-BC) total severity score (1A), Childhood Behavior Checklist (CBCL) total t-score (1B), and Schizophrenia Spectrum Disorder – Regional Vulnerability Indices (SSD-RVIs) (1C) across the cognitive subgroups. Error bars represent standard errors.

At baseline, there was no significant correlation between SSD-RVIs and PQ-BC severity scores (*r* = - 0.036, *p* = 0.3, *q* = 0.3). There was a weak trend-level correlation between SSD-RVIs and NIHTB age-corrected total composite scores (*r* = -0.060, *p* = 0.092, *q* = 0.138), as well as with CBCL total t-scores (*r* = -0.066, *p* = 0.063, *q* = 0.138). However, neither correlation survived FDR correction.

### The effects of age and the interaction between age and cognitive subgroups in the longitudinal data

There was a significant effect of age on PQ-BC total severity and CBCL total t scores during the follow-up period in the whole FHR group. Both PQ-BC (β = - 1.14, 95% CI [-1.45, -0.83], *t* = -7.232, *q* < 0.001, *d* = -0.13) and CBCL (β = - 0.40, 95% CI [-0.64,-0.14], *t* = -3.126, *q* = 0.003, *d* = -0.06) scores significantly decreased with age over time. SSD-RVIs remained stable with age (*q* = 0.8).

There were no significant age * cognitive subgroup interactions on PQ-BC total severity scores, CBCL total t scores, or SSD RVIs (*q’s* > 0.05), indicating no differences in how these outcomes change with age across the three cognitive subgroups over the follow-up period (Figure 2).

**Figure 2.**
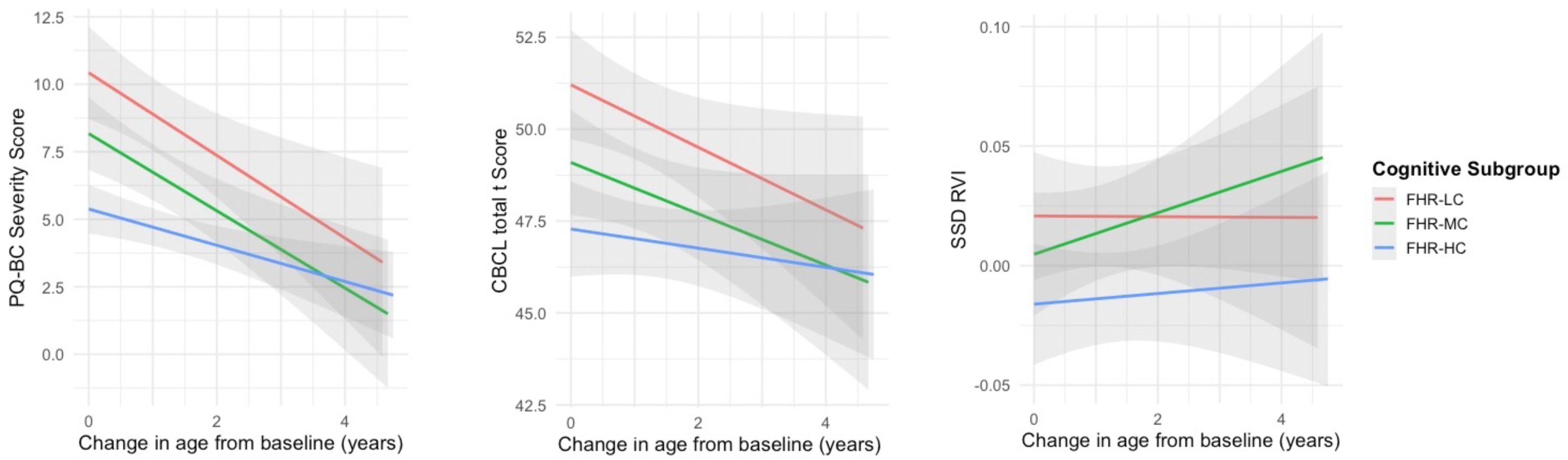
Line graphs with shaded confidence intervals, displaying the change of Prodromal Questionnaire-Brief Child Version (PQ-BC) severity score, Childhood Behavior Checklist (CBCL) total t- score, and Schizophrenia Spectrum Disorder – Regional Vulnerability Indices (SSD-RVIs) with age during the follow-up period (baseline, year 2, year 4) across the cognitive subgroups. FHR-LC: Family High Risk – Low Cognition, FHR-MC: Family High Risk – Moderate Cognition, FHR-HC: Family High Risk – High Cognition.

### Results of the sensitivity analysis

Please see **Supplementary Material** for full results of the sensitivity analysis and a discussion of these results. The results of the complete case analysis demonstrated trend-level higher PQ-BC total severity scores (*p* = 0.079, *q* = 0.25, *d* = 0.84) and higher SSD-RVI scores in FHR-LC compared to FHR-HC (*p* = 0.016, *q* = 0.147, *d* = 0.48) at baseline, not surviving FDR correction. CBCL total t scores were not significantly different across the three groups at baseline. The effect of age on PQ-BC total severity and CBCL total t scores during the follow-up period remained significant in participants with complete data (PQ-BC, *q* = 0.007, *d* = -0.13; CBCL, *q* < 0.001, *d* = -0.07). SSD-RVIs remained stable with age (*q* = 0.98). There were no significant age * cognitive subgroup interactions on PQ-BC total severity scores, CBCL total t scores, or SSD RVIs (*q’s* > 0.05) also in the complete case analysis. Overall, results from the sensitivity analyses were generally consistent with our primary analysis, with the exception of baseline group differences in CBCL total t scores, supporting the robustness of our findings.

## DISCUSSION

This study demonstrated that, among 9-10 year-old children with a family history of psychosis, FHR children with low cognitive performance (0-33 percentile) reported more severe psychotic-like experiences and had more childhood psychiatric symptoms compared to FHR children with high cognitive performance (66-100 percentile). FHR children with low cognitive performance also demonstrated more similar brain patterns to those observed in adults with schizophrenia at a trend-level significance. PLEs and general psychopathology scores decreased with age during the 2- and 4-year follow-up periods in all FHR participants, while SSD-RVIs remained stable. There was no interaction between the effects of cognitive subgroup membership and age on outcome measures (PQ-BC, CBCL, SSD-RVI) longitudinally, which suggests that the group differences in these measures remained stable with age during the follow up period. A sensitivity analysis, including only participants with complete BL, Y2, an Y4 data, upheld these findings, except for baseline group differences in the CBCL total t scores, further supporting the robustness of these results.

Our findings suggest that, among FHR children, those with low cognitive performance are at an heightened risk for poorer mental health outcomes compared to moderate to high cognitive performers. These results align with results from the Danish High Risk and Resilience Study, VIA-7 and VIA-11 arms (7). Knudsen et al. demonstrated that, among FHR children, the severely-moderately impaired cognitive subgroup had higher CBCL scores and lower global functioning compared to mildly-impaired and above average cognitive subgroups both at ages 7 and 11. In a different FHR cohort of children ages between 6-17, Valli et al. demonstrated higher overall pathology and decreased general functioning in the impaired cognitive subgroup compared to the intact and intermediate cognitive subgroups (17). Our findings add to these results providing evidence that the cognitively impaired FHR subgroup also reports more psychotic-like experiences and has brain patterns more similar to those observed in adults with schizophrenia spectrum disorders. These results are also closely in line with the strong evidence demonstrating lower premorbid cognitive functioning in individuals who later develop psychotic disorders (18,19). Taken together, these findings suggest that FHR children with low cognitive performance represent a well-defined and early identifiable risk group for future psychotic disorders.

We demonstrated that cognitive subgroup differences in outcome measures remained stable with age during the 4-year follow up period, although PQ-BC and CBCL scores decreased with age in the whole FHR group. These results are not in line with our hypotheses. We hypothesized a steeper increase in clinical symptoms and SSD-RVIs with age in the FHR-LC group. However, these findings also show parallels with the Danish High Risk and Resilience Study (7). Knudsen et al. demonstrated that CBCL scores decreased with age from age 7 to 11 in all three cognitive subgroups, despite remaining group differences. This is also consistent with the normative development of child and adolescent problem behavior. Using a birth cohort sample of 2600 children, Bangers et al. demonstrated that total CBCL scores decreased with time from age 4 to 18, with distinct trajectories for internalizing and externalizing symptoms (20). Studies show a similar trend for PQ-B scores. Pelizza et al. showed a negative correlation with age in PQ-B scores in a sample of help-seekers aged 13-35, with younger adolescents (age <15) showing higher perceptual abnormality scores in the Comprehensive Assessment of At-Risk Mental States (CAARMS)(21). We hypothesize that there may be a curvilinear relationship between age and clinical symptoms in FHR children. Specifically, we predict that CBCL and PQ-BC scores will decrease during early to mid-adolescence, following a similar developmental trend observed in healthy children with no family history of psychosis. However, we expect a subsequent increase in psychotic-like symptoms during late adolescence and early adulthood, particularly in the FHR-LC subgroup, coinciding with the critical period when psychosis prodrome typically occurs. It is also possible that these measures may not be predictive of future psychosis risk in children and adolescents. The ABCD study will follow participants until age 18-20; however, data collection has not been completed. We plan to test these hypotheses in future data releases.

Our results further contribute to the literature by demonstrating that the low cognition FHR subgroup also exhibits structural brain patterns more similar to those observed in adults with schizophrenia spectrum disorders. Previously, Kochunov et al. demonstrated that there were no differences in SSD-RVIs between children with and without an ancestral history of SSD in the ABCD baseline dataset, despite a correlation between SSD-RVIs and family history loadings (10). Our findings showing a trend-level difference in SSD-RVIs among FHR children with low and high cognitive performance indicates that SSD-RVIs may function better as a marker for stratifying risk within high-risk populations rather than as a marker of psychosis risk in general populations. Similarly, Karcher et al. showed that, in comparison to children with low distressing PLE’s (i.e. healthy control group), only children with persistent and distressing PLEs had increased SSD-RVIs in the ABCD dataset (9). Children with persistent nondistressing, transient distressing, and transient nondistressing PLEs exhibited no significant differences in SSD-RVIs compared to healthy controls. Together, these suggest that SSD-RVIs may have higher specificity, but low sensitivity when applied broadly as a marker of psychosis risk. Overall, these studies indicate that structural brain patterns resembling those observed in adults with schizophrenia emerge as early as ages 9-10 in children at increased risk for psychosis, highlighting the critical importance of implementing preventive interventions during early childhood in this vulnerable population.

It is important to highlight that the FHR-LC group composed of more children from minority racial and ethnic backgrounds (73%) compared to the FHR-HC group (39%). This stark difference suggests that FHR children from minority backgrounds are more likely to lag behind their peers in cognitive development. These findings underscore the apparent impact of equity issues on brain development (22,23), as systemic barriers such as socioeconomic disparities, access to resources, and educational opportunities disproportionately affect minority populations. In FHR populations, this disparity is likely even more pronounced, given that equity issues also influence the affected relative’s access to care and effective treatment, creating a compounded negative impact on the child’s cognitive development and mental health outcomes. We did not control for socioeconomic status in our analyses because we consider it a contributing factor to the outcomes we examined in FHR children, rather than a confounding factor. Our findings indicate that FHR children from racial and ethnic minority backgrounds represent another higher-risk subgroup within the FHR population, identifiable from birth.

There is limited evidence on effective preventive interventions for children at risk of psychosis (24–26), but this gap arises partly from the study designs typically used in psychiatric research. Randomized clinical trials (RCTs) pose significant practical and ethical concerns when applied to preventive approaches in children targeting neurodevelopmental disorders. We propose reframing a developmental question: “Do preventive interventions that support healthy brain development improve brain and mental health metrics in children at risk for severe mental illness?” Existing research on neurodevelopment already provides extensive evidence on factors that promote healthy brain development. These include adequate prenatal care, optimal nutrition, regular physical activity, positive parent-child interactions, access to early education, adequate sleep, and protection from adverse childhood experiences and substance use (27–32). Although these factors are not specific to mitigating psychosis risk and are essential for every child, prioritizing limited resources to address them is particularly justified for vulnerable FHR children. There is a need for family-based preventive programs catered for FHR children that address these factors through parental training, family therapy, skills-building and cognitive trainings, early risk assessment, and interventions for early risk signs (25,33). Implementing such programs has the potential to not only reduce the risk of psychosis but also improve overall developmental outcomes and quality of life for FHR children and their families.

A major limitation of this study is that the FHR status is not determined based on clinical interviews with affected relatives. Family history status was determined based on questions in the ABCD baseline assessment, which asked whether relatives had experiences such as seeing visions or hearing voices, or thinking of people were spying on them or plotting against them. As these questions are not specific for psychotic disorders, there are likely false positives among the FHR children in our sample. Additionally, among the true positive FHR cases, we don’t know the affected relatives’ diagnoses or the severity of their disorders, which could affect our outcome measures. Another limitation is the partially released 4-year follow-up data and attrition during the follow-up period (74% smaller sample size in Y4 compared to BL). The sample size reduction is even higher for the FHR-LC group (78%) compared to the FHR-HC group (65%). The disproportionate and smaller group sample sizes in the Y4 data (FHR-LC, n=50; FHR-MC, n=54; FHR-HC, n=96) may have reduced the statistical power of our study to detect longitudinal changes over the 4 years, increasing the risk of Type II errors. Despite this, the results of a sensitivity analysis only including participants with complete BL, Y2, and Y4 data were largely consistent with the findings of our primary analyses. The sample size precluded analyses of sex differences, which are likely present in both clinical and brain development trajectories. We also did not examine the existing mental health diagnoses in our FHR cohort, as this analysis was beyond the scope of our hypotheses and planned analyses. Additionally, these findings are specific to FHR children. Given that our primary goal was to identify risk subgroups within FHR children, we did not include a sample with low cognition without FHR.

In conclusion, among 9-10-year-old children with a family history of psychosis within a large developmental cohort, we found that those with lower cognitive performance exhibited more severe psychotic-like experiences, greater general childhood psychopathology, and structural brain patterns more similar to those observed in adults with schizophrenia, compared to their peers with higher cognitive performance. Notably, these differences persisted over the four-year follow-up period. These findings indicate that FHR children with low cognitive performance represent an easily identifiable subgroup at heightened developmental risk for adverse mental health outcomes. This subgroup can be detected early in life, providing a crucial window for preventive interventions aimed at supporting healthy brain development and mitigating long-term modifiable risks.

## Data Availability

Data used in the preparation of this article were obtained from the Adolescent Brain Cognitive Development (ABCD) Study (https://abcdstudy.org), held in the NIMH Data Archive (NDA). The ABCD data used in this report came from https://nda.nih.gov/study.html?id=2313.

## Acknowledgments

This work has been supported by the Brain and Behavior Research Foundation Young Investigator Grant (PI: Dr. Türközer). Additionally, Dr. Türközer is supported by Harvard Medical School’s Dupont Warren and Livingston Fellowships. Dr. Karcher is supported by the National Institute of Mental Health (NIMH) (K23 MH121792). Dr. Clauss is supported by the Maryland Psychiatric Research Center and the Chen Institute Mass General Neuroscience Transformative Scholar Award. Dr. Roffman is supported by NIMH (R01MH124694) and the Mass General Early Brain Development Initiative. Dr. Öngür is supported by NIMH (P50MH115846).

Data used in the preparation of this article were obtained from the Adolescent Brain Cognitive Development (ABCD) Study (https://abcdstudy.org), held in the NIMH Data Archive (NDA). This is a multisite, longitudinal study designed to recruit more than 10,000 children ages 9–10 and follow them over 10 years into early adulthood. The ABCD Study is supported by the National Institutes of Health and additional federal partners under award numbers U01DA041022, U01DA041025, U01DA041028, U01DA041048, U01DA041089, U01DA041093, U01DA041106, U01DA041117, U01DA041120, U01DA041134, U01DA041148, U01DA041156, U01DA041174, U24DA041123, and U24DA041147. A full list of supporters is available at https://abcdstudy.org/nih-collaborators. A listing of participating sites and a complete listing of the study investigators can be found at https://abcdstudy.org/principal-investigators.html. ABCD consortium investigators designed and implemented the study and/or provided data but did not necessarily participate in analysis or writing of this report. This manuscript reflects the views of the authors and may not reflect the opinions or views of the NIH or ABCD consortium investigators. The ABCD data repository grows and changes over time. The ABCD data used in this report came from https://nda.nih.gov/study.html?id=2313.

