## Supplement for "Psychosis-linked Symptoms and Structural Brain Patterns in Cognitive Subgroups among Familial High-Risk Children in the ABCD Study"

**Supplementary Material**

***The change in cognitive subtests with age over time across the cognitive subgroups***

We utilized linear mixed-effects models using the lme4 package in R to examine how cognitive subtest performances changed with age over time across the three cognitive subgroups. The cognitive subgroups were determined based on baseline NIH Toolbox total age-corrected scores: FHR-LC: Familial high risk – low cognitive performance (0-33%), FHR-MC: Familial high risk – moderate cognitive performance (33-66%), FHR-HC: Familial high risk – high cognitive performance (66-100%). Age corrected scores from five NIH Toolbox subtests, which were available in the BL, Y2, and Y4 data, were included in the longitudinal analysis: picture vocabulary, flanker inhibitory control, pattern comparison processing speed, picture sequence memory, and oral reading recognition. We couldn’t include the total scores in this analysis, as total scores were not available in the Y2 and Y4 data.

We first evaluated the fixed effect of age longitudinally in age-corrected subtest scores in the whole FHR group to assess their change with age over time. We then examined the interaction between age and cognitive subgroups on cognitive subtest scores longitudinally. Subject ID was added as a random factor. Family ID and site were also added as random effects to account for clustering and site-specific effects. Covariates included sex and race-ethnicity. The false discovery rate (FDR) approach was used to correct for multiple testing (*q* value).

*Results*

In the whole FHR group, picture vocabulary scores decreased with age over time (β  = -1.44, 95% CI [-1.79, -1.096], *t* = -8.135, q<0.001, *d* = -0.16). Pattern comparison processing speed (β  = 5.71, 95% CI [5.12, 6.13], *t* = 18.794, q<0.001, *d* = 0.36), picture sequence memory (β  = 1.84, 95% CI [1.36, 2.32], *t* = 7.55, q<0.001, *d* = 0.14), and oral reading (β  = 0.38, 95% CI [0.029, 0.75], *t* = 2.12, q=0.042, *d* = 0.04) scores increased with age over time. Flanker inhibitory control scores did not change over time (q>0.05). There were no significant cognitive subgroup * age interactions on any cognitive subtests, except for picture sequence memory score. There was a significant age by group interaction on picture vocabulary score for the FHR-HC and FHR-LC groups, suggesting a more steep decline with age in the FHR-HC group compared to FHR-LC (β  = -1.40, 95% CI [-2.23, -0.57], *t* = -3.32, q=0.013, *d* = -0.15) (Figure S1).

***Results of the sensitivity analysis***

Following the analyses on the whole dataset, we conducted a sensitivity analysis by performing a complete case analysis, including only participants with complete BL, Y2, an Y4 data (N=175, n_FHR-LC_=42, n_FHR-MC_=48, n_FHR-HC_=85) using the same methods described above. To assess the robustness of findings, we compared results from the complete case analysis with those obtained using the entire dataset.

A complete case analysis demonstrated that, at baseline, FHR-LC had higher PQ-BC total severity scores compared to FHR-HC at trend level significance, which did not survive FDR correction (β  = 4.12, 95% CI [-0.440, 8.672], *t* = 1.77, *p* = 0.079, *q* = 0.25, *d* = 0.84). CBCL total t scores were not significantly different across the three groups. FHR-LC had higher SSD-RVI scores compared to FHR-HC, however, this difference did not survive FDR correction (β = 0.099, 95% CI [0.019, 0.179], *t* = 2.42, *p* = 0.016, *q* = 0.147, *d* = 0.48).

The effect of age on PQ-BC total severity and CBCL total t scores during the follow-up period remained significant in participants with complete data. Both PQ-BC (β  = - 1.07, 95% CI [-1.49, -0.66], *t* = -5.131, *q* = 0.007, *d* = -0.13) and CBCL (β  = - 0.49, 95% CI [-0.83,-0.15], *t* = -2.831, *q* < 0.001, *d* = -0.07) scores significantly decreased with age over time. SSD-RVIs remained stable with age (*q* = 0.98).

There were no significant age * cognitive subgroup interactions on PQ-BC total severity scores, CBCL total t scores, or SSD RVIs (*q’s* > 0.05) also in the complete case analysis.

*Discussion*

This sensitivity analysis demonstrates that, at baseline, the higher PQ-B severity scores and SSD-RVI scores in the FHR-LC group compared to the FHR-HC group were upheld, despite the disproportionally reduced sample size in the FHR-LC group and the lower power of the analysis. While these findings did not survive FDR correction in the smaller sample, the direction of effects and substantial effect sizes suggest consistency with the primary analysis. However, the non-significant results for CBCL total t scores in the complete case analysis indicate that these findings are more sensitive to sample size and missing data.

The significant effect of age on PQ-BC total severity and CBCL total t scores was consistent across the entire dataset and the complete case analysis. SSD-RVI scores remained stable with age, and no significant age × cognitive subgroup interactions were observed for any of the three outcomes in either analyses, suggesting the reliability of the findings despite the smaller sample size in the sensitivity analysis.

Overall, the results of the sensitivity analysis were generally consistent with our primary analysis using the entire dataset, with the exception of baseline group differences in the CBCL total t scores. These results suggest that our analyses had adequate power to detect increased severity of psychosis-linked symptoms and brain patterns amongst individuals who have not yet completed 4-year follow-up or were lost to follow-up.

It is also worth noting that children who complete the assessment at the tail end of the assessment period and those who are lost to follow-up may have on average higher levels of psychopathology. This is consistent with the disproportionately smaller sample sizes in the FHR-LC and FHR-MC groups in the Y4 data (FHR-LC, n=50; FHR-MC, n=54; FHR-HC, n=96). This may have reduced the statistical power of our study to detect longitudinal changes over the 4 years particularly in the FHR-LC and FHR-MC groups, increasing the risk of Type II errors.

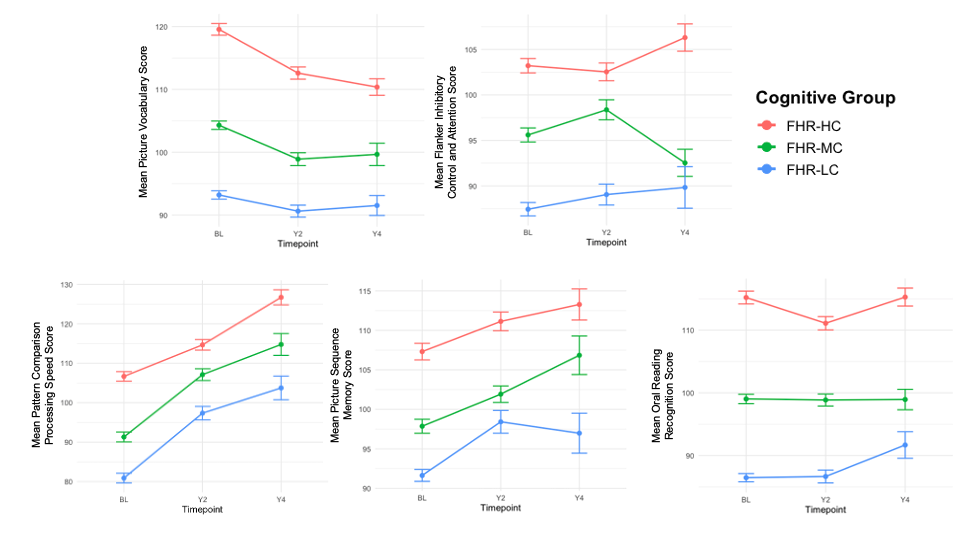

**Figure S1.** NIH Toolbox subtest age-corrected scores that are available in BL, Y2, and Y4 data across the three cognitive subgroups. Cognitive subgroups were determined based on **baseline** total age corrected scores. FHR-LC: Familial high risk – low cognitive performance (0-33%), FHR-MC: Familial high risk – moderate cognitive performance (33-66%), FHR-HC: Familial high risk – high cognitive performance (66-100%). Error bars represent standard errors.
